# Transmission potential of COVID-19 in South Korea

**DOI:** 10.1101/2020.02.27.20028829

**Authors:** Eunha Shim, Amna Tariq, Wongyeong Choi, Yiseul Lee, Gerardo Chowell

**Author notes:** **Corresponding author:** Eunha Shim, **Corresponding author email:**. **Conflict of interest**: None.

## Abstract

Since the first identified individual of 2019 novel coronavirus (COVID-19) infection on Jan 20, 2020 in South Korea, the number of confirmed cases rapidly increased. As of Feb 26, 2020, 1,261 cases of COVID-19 including 12 deaths were confirmed in South Korea. Using the incidence data of COVID-19, we estimate the reproduction number at 1.5 (95% CI: 1.4-1.6), which indicates sustained transmission and support the implementation of social distancing measures to rapidly control the outbreak.

---

A novel coronavirus (SARS-CoV-2) that emerged out of the city of Wuhan, China in December 2019 has already demonstrated its potential to generate explosive outbreaks in confined settings and cross borders following human mobility patterns (1). While COVID-19 frequently induces mild symptoms common to other respiratory infections, it has also exhibited an ability to generate severe disease among certain groups including older populations and individuals with underlying health issues such as cardiovascular disease and diabetes (2). Nevertheless, a clear picture of the epidemiology of this novel coronavirus is still being elucidated.

The number of cases of COVID-19 in the province of Hubei, the disease epicenter, quickly climbed following an exponential growth trend. The total number of COVID-19 cases is at 81,109 including 2718 deaths in China as of February 26, 2020 (3). Fortunately, by February 15, 2020 the daily number of new reported cases in China started to decline across the country although Hubei Province is still reporting hundreds of cases per day as of February 26^th^, 2020 (3). While the epidemic continues to decline in China, more than 2,900 COVID-19 cases have been reported in 37 countries outside of China including South Korea, Japan, Italy, Iran, Singapore, U.S., and Thailand (3). In particular, South Korea has quickly become the second hardest hit country with COVID-19, with steadily increasing number of cases over the last few days. Hence, it is crucial to monitor the progression of these outbreaks and assess the effects of various public health measures including the social distancing measures in real time.

The first case in South Korea was identified on January 20, 2020 followed by the detection of one or two cases on average in the subsequent days. However, the number of confirmed cases of SARS-CoV-2 infection started to increase rapidly on February 19, 2020 with a total of 1,261 confirmed COVID-19 cases including twelve deaths reported as of February 26, 2020 according to the Korea Centers for Disease Control and Prevention (KCDC) (4). The epicenter of the South Korean COVID-19 outbreak has been identified in Daegu, a city of 2.5 million people, approximately 150 miles South East of Seoul. The rapid spread of COVID-19 in South Korea has been attributed one case linked to a superspreading event that has led to at least 40 secondary cases stemming from church services in the city of Daegu (5, 6). This has led to sustained transmission chains of COVID-19, with about 52% of the cases associated with the church cluster in Daegu (7). Moreover, three other clusters have been reported including one set in Chundo Daenam hospital in Chungdo-gun, Gyeongsanggbuk-do (114 cases), one Pilgrimage to Israel cluster in Gyeongsanggbuk-do (31 cases) and a cluster A composed of one imported and 11 local cases in Seoul. These few clusters have become the major driving force of the infection. A total of 33 cases were imported while the three major clusters are composed of local cases as described in Supplementary Table S1.

Transmission of SARS-CoV-2 in Korea has been exacerbated by amplified transmission in confined settings including a hospital and a church in the city of Daegu. The hospital-based outbreak alone involves 114 individuals including 9 hospital staff (8), which is reminiscent of past outbreaks of SARS and MERS (9). To respond to the mounting number of cases of COVID-19, the Korean government has raised the COVID-19 alert level to the highest (Level 4) to facilitate the implementation of social distancing measures including enhanced infection control measures in hospitals, restricting public transportation, cancelling social events, and delaying the start of school activities in order to contain the outbreak (10).

The effective reproduction number (*R*_*t*_) quantifies the time-dependent transmission potential of an infectious diseases outbreak. This key epidemiological parameter tracks the average number of secondary cases generated per case as the outbreak progresses over time. Steady values of *R*_*t*_ above 1 indicate sustained disease transmission, whereas values of *R*_*t*_ <1 do not support sustained transmission and the number of new cases is expected to follow a declining trend. In this report, using a mathematical model parameterized with cases series of the COVID-19 outbreak in Korea, we investigate the transmission potential of COVID-19 in Korea using data of local and imported cases up until February 26, 2020.

## Methods

### Data

We obtained the daily series of confirmed cases of COVID-19 in South Korea from January 20, 2020 to February, 26, 2020 that are publicly available from the Korea Centers for Disease Control and Prevention (KCDC) (4). Our data includes the dates of reporting for all confirmed cases, the dates of symptom onsets for the first 28 reported cases, and whether the case is autochthonous (local transmission) or imported. We also summarize the case clusters comprising one or more cases according to the source of infection according to the field investigations conducted by the KCDC (4). Accordingly, four major clusters were identified.

### Imputing the date of onset

To estimate the growth rate of the epidemic, it is ideal to characterize the epidemic curve according to dates of symptoms onset rather than according to dates of reporting. For the COVID-19 data in Korea, the symptom onset dates are available for only the first 28 reported cases. Moreover, all of the dates of symptoms onset are available for the imported cases. Therefore, we utilize this empirical distribution of reporting delays from the onset to diagnosis to impute the missing dates of onset for the remainder of the cases with missing data. For this purpose, we reconstruct 300 epidemic curves by dates of symptoms onset from which we derived a mean incidence curve of local case incidence and drop the last three data points from the analysis to adjust for reporting delays in our real-time analysis (11).

### Estimate of reproduction number from daily case incidence

We assess the effective reproduction number, *R*_*t*_, which quantifies the time dependent variations in the average number of secondary cases generated per case during the course of an outbreak due to intrinsic factors (decline in susceptible individuals) and extrinsic factors (behavior changes, cultural factors, and the implementation of public health measures) (9, 12, 13). Using the Korean incidence curves for imported and local cases, we estimate the evolution of *R*_*t*_ for COVID-19 in Korea. First, we characterize daily local case incidence using the generalized growth model (GGM) (14). This model characterizes the growth profile via two parameters: the growth rate parameter (*r*) and the scaling of the growth rate parameter (*p*). The model captures diverse epidemic profiles ranging from constant incidence (*p* = 0), sub-exponential or polynomial growth (0 < *p* < 1), and exponential growth (*p* = 1) (14). The generation interval is assumed to follow a gamma distribution with a mean of 4.41 days and a standard deviation of 3.17 days (15, 16).

Next, to estimate the most recent estimate of *R*_*t*_, we simulate the progression of incident cases from GGM, and apply the discretized probability distribution (*ρ*_*i*_) of the generation interval using the renewal equation (17-19) given by 

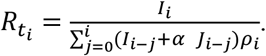

In the renewal equation we denote the local incidence at calendar time *t*_*i*_ by *I*_*i*_, and the raw incidence of imported cases at calendar time *t*_*i*_ by *J*_*i*_. The parameter 0 ≤ *α* ≤ 1 quantifies the relative contribution of imported cases to the secondary disease transmission (20). The denominator represents the total number of cases that contribute to the incidence cases at time *t*_*i*_. Next, we estimate *R*_*t*_ for 300 simulated curves assuming a Poisson error structure to derive the uncertainty bounds around the curve of *R*_*t*_ (21).

## Results

### Reconstructed incidence of COVID-19

The reconstructed daily incidence curve of COVID-19 after the imputing the onset dates for the Korean cases is shown in Figure 1. Between January 20 and February 18, 2020 an average of 2 new cases were reported each day, whereas between February 19-26, 2020, 153 new cases were reported each day on average.

**Figure 1:**
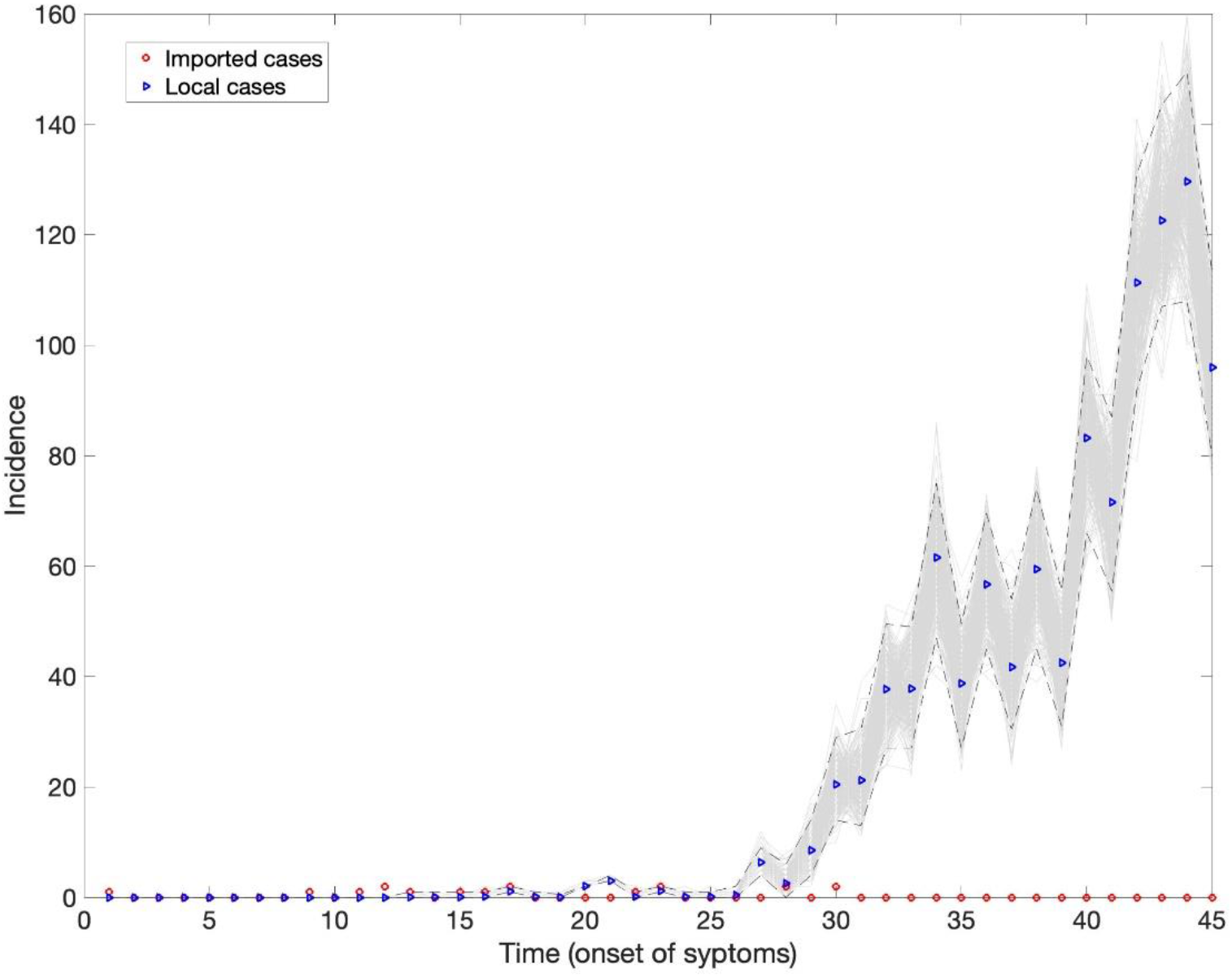
Reconstructed epidemic curve for the local Korean COVID-19 cases by the dates of onset. The blue triangles represent the local cases, red triangles represent the imported cases and the gray curves correspond to the uncertainty in the local cases because of missing onset dates.

### Effective reproduction number (R_t_) from daily case incidence

Under the empirical reporting delay distribution from Korean cases with available dates of onset, the intrinsic growth rate (*r*) was estimated at 0.6 (95% CI: 0.5, 0.7) and the scaling of growth parameter (*p*) was estimated at 0.76 (95% CI: 0.73, 0.79), indicating sub-exponential growth dynamics of COVID-19 in Korea (Figure 2, Table 1). The mean reproduction number *R*_*t*_ was estimated at 1.5 (95% CI: 1.4, 1.6). Our estimates of *R*_*t*_ are not sensitive to the changes to the parameter that modulates the contribution of the imported cases to transmission (*α*).

**Table 1:** Mean estimates and the corresponding 95% confidence intervals for the effective reproduction number, growth rate and the scaling of growth parameter during the early growth phase as of February 26, 2020.

| Parameters | Estimated values |
| --- | --- |
| **Reproduction number** | 1.5 (95% CI:1.4,1.6) |
| **Growth rate, $r$** | 0.6 (95%CI:0.5,0.7) |
| **Scaling of growth parameter, $p$** | 0.76 (95%CI:0.73,0.79) |

**Figure 2:**
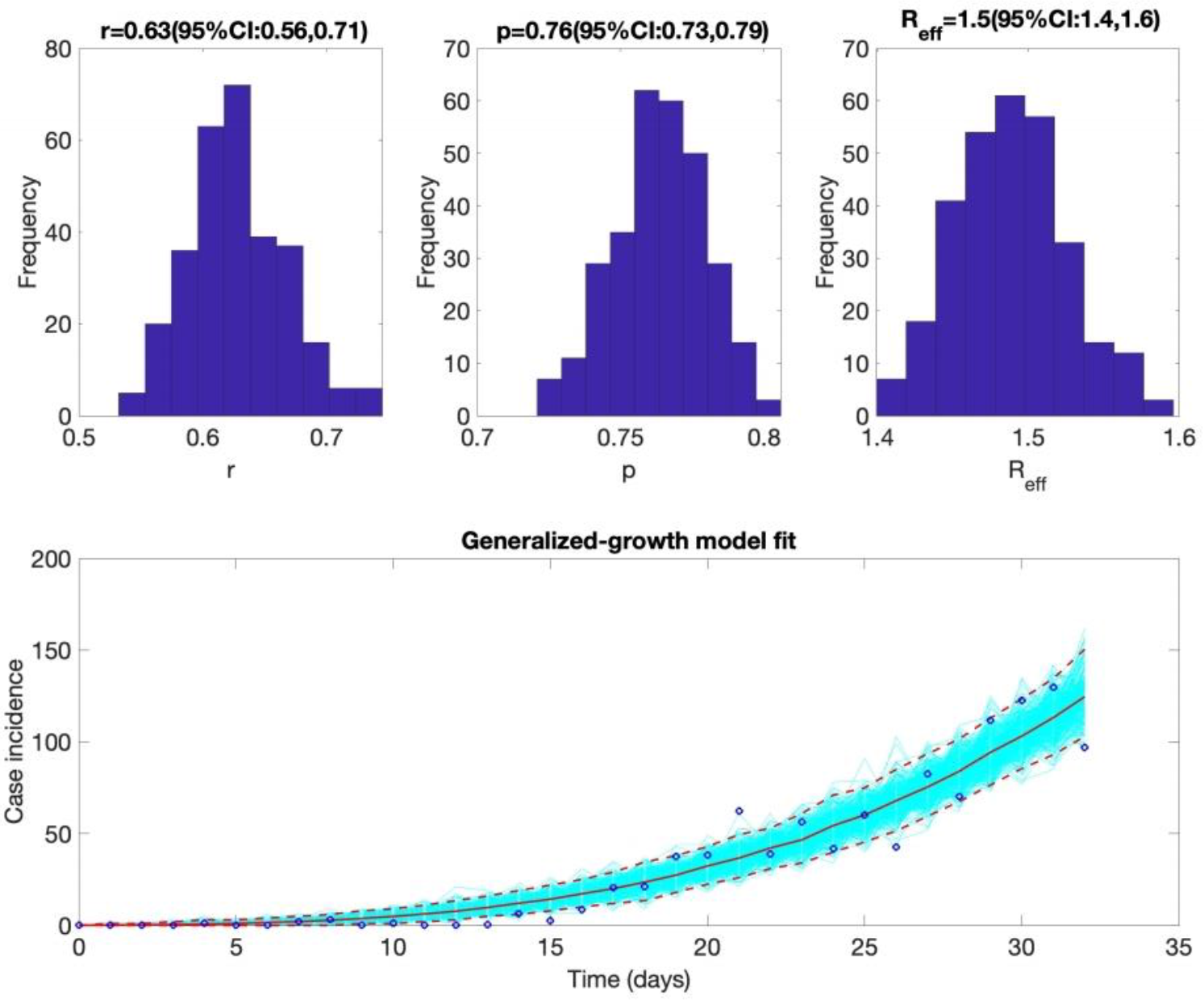
The mean reproduction number with 95% CI estimated by adjusting for the imported cases with *α*=0.15. Estimates for growth rate (*r*) and the scaling of the growth rate parameter (*p*) are also provided. The plot at the bottom depicts the fit of the Generalized Growth Model to the Korean data assuming Poisson error structure as of February 26, 2020.

### Transmission clusters

Spatial distribution of the Korean clusters is shown in Figure 3 and the characteristics of each cluster are presented in Table 1.

**Figure 3:**
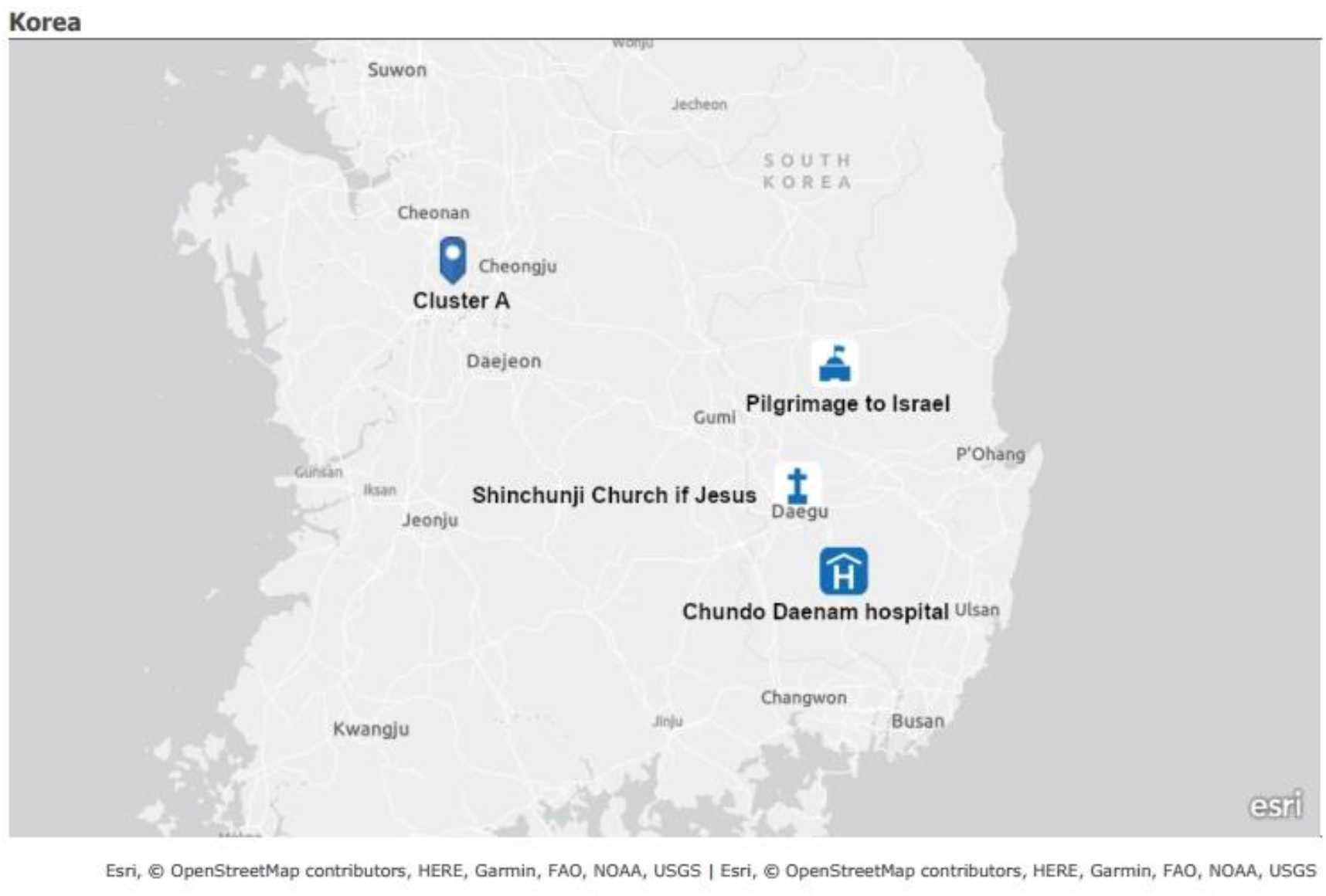
Map depicting the spatial distribution of the four largest clusters of COVID-19 in Korea as of February 26, 2020.

### Shincheonji Church of Jesus cluster

As of February 26, 2020, 597 (or 47%) of the country’s 1261 confirmed cases of COVID-19 are linked to this cluster, according to the KCDC (4). This largest cluster is associated with Shincheonji Church of Jesus, with the first case (the 31^st^ patient in the country) confirmed on Feb. 18. It is unclear how this case contracted the illness, as she does not present a recent history of travel or contact with another infected patient. However, this case attended Shincheonji Church of Jesus in Daegu four times before and after becoming a symptomatic case of COVID-19, and visited the hospital in Cheongdo after a car accident. According to the KCDC, the patient had contact with 166 people mostly at Shincheonji Church and the hospital in Cheongdo, who now placed themselves into self-quarantine. The Shincheonji church of Jesus has temporarily closed its facilities and halted the church activities as of February 18, 2020.

### Pilgrimage tour to Israel related

A total of 31 infected cases belonging to a group of 39 Catholic pilgrims who visited Israel between February 8, 2020 and February 16, 2020 were confirmed for COVID-19 (22). Amongst these, 11 individuals were diagnosed on February 17, 2020, while 20 others were confirmed positive between February 21-25, 2020 and quarantined immediately. Of the 31 infected pilgrims, 19 belong to the Euiseong County, North Gyeongsang Province, while one patient, a tour guide belongs to Seoul. Health authorities have traced 170 contacts from these 29 cases, raising concerns about the potential risk of secondary infections.

### Chungdo Daenam hospital cluster

This cluster comprised of 114 local cases and five deaths is associated with Chungdo Daenam hospital, where South Korea’s first coronavirus associated case fatality occurred. Of the 114 cases, 92 were confirmed on Feb 22, 2020 (4). A 63-year-old man who died of pneumonia at the hospital on Feb 19 was posthumously tested positive for COVID-19. On Feb 21, another patient at Daenam Hospital died from COVID-19, followed by another death on Feb 23. The confirmed cases were mainly from psychiatric ward and includes nine medical staffs. The exact route of the infection is not yet known.

### Cluster A

This cluster is composed of one imported case and 12 local cases, and was identified with its first case (the 3rd patient in the country) confirmed on Jan 26, 2020. Its first case is suspected to be an imported case from Wuhan, China, transmitting SARS-CoV-2 to the 6^th^ and 28^th^ confirmed cases in Korea, which resulted in secondary cases. No further cases have been added in this cluster since February 21, 2020.

## Discussion

This is the first study to report estimates of the transmission potential of COVID-19 in Korea based on the trajectory of the epidemic, which was reconstructed by using the dates of onset of the first reported cases in Korea. The estimates of *R* clearly indicate sustained transmission of the novel coronavirus in Korea. Moreover, the imported cases contribute little to the secondary disease transmission in Korea, as majority of these cases occurred in the early phase of the epidemic, with the most recent imported case reported on February 9, 2020. These findings support the range of social distancing interventions that the Korean government put in place in order to bring the outbreak under control as soon as possible.

Our estimates of the reproduction number can be compared with earlier estimates reported for the epidemic in China where the estimates of *R* lie in the range 2-7.1 (1, 23-29). Moreover, the mean *R* reached values as high as ∼11 for the outbreak that unfolded aboard the Princess Cruises Ship during January-February 2020 (30). In contrast, a recent study on Singapore’s COVID-19 transmission dynamics reported lower estimates for *R*_*t*_ (1.1, 95% CI: 1.1, 1.3) as of February 19^th^, 2020, reflecting a significant impact of the control interventions that have been implemented in Singapore (31). The estimates of the scaling of growth parameter (*p*) in our study indicate sub-exponential growth dynamics of COVID-19 in Korea. This aligns well with the sub-exponential growth patterns of COVID-19 in Singapore and all Chinese provinces except Hubei (31, 32).

Since the first COVID-19 case was reported on January 20, 2020, the epidemic’s trajectory showed a rapid upturn until February 18, 2020, when a super spreader (Case 31) was identified in the Shincheonji Church of Jesus in Daegu cluster. Since then, Korea’s confirmed cases have risen tremendously. In fact, 80 percent of confirmed cases are linked to the two clusters of infections, i.e. Shincheonji Church of Jesus in Daegu and Chungdo Daenam hospital (33). Such superspreading events have been reported for the 2015 MERS outbreak in South Korea (34). Amplification of MERS in the hospital setting has been associated with diagnostic delays, which increase the window of opportunity for the generation of secondary cases (9). This underscores the need for rapid testing, case detection and active contact tracing to isolate infectious individuals.

Beyond Korea, substantial COVID-19 transmission has been reported in Singapore, Iran, Italy, Germany and aboard the Diamond cruise ship (35, 36). While the cluster A and the Chungdo Daenam hospital cluster in Korea seem to have stabilized, the other two clusters are likely to keep growing in the short term. Public health authorities are currently focused on containing the outbreak in the city of Daegu, the epicenter of the outbreak and North Gyeongsang Province where active contact tracing is being conducted. Nation-wide preventative measures are expected to reduce the community transmission and ultimately bring *R*_*t*_ below one.

## Conclusion

This is the first study to estimate the transmission potential of COVID-19 in Korea. Our current findings suggest sustained disease transmission in the region. Our estimates support the implementation of a wide array of social distancing measures to rapidly contain the outbreak in Korea.

## Data Availability

We obtained the daily series of confirmed cases of COVID-19 in South Korea from January 20, 2020 to February, 26, 2020 that are publicly available from the Korea Centers for Disease Control and Prevention (KCDC)

https://www.cdc.go.kr/board/board.es?mid=a30402000000&bid=0030

